# Pulmonary Vascular Compromise is Associated with Survival in Pediatric Pulmonary Hypertension: A New Computational Model

**DOI:** 10.1101/2025.03.25.25324654

**Authors:** Maria Niccum, Catherine M. Avitabile, Dana Albizem, Heather Meluskey, Christopher Penney, Brian D. Hanna, Michael L. O’Byrne, Zoheir Bshouty, David B. Frank

**Affiliations:** Department of Pediatrics, Division of Cardiology, Perelman School of Medicine at the University of Pennsylvania, Children’s Hospital of Philadelphia, Philadelphia, PA 19104; Division of Biomedical and Health Informatics, Children’s Hospital of Philadelphia, Philadelphia, PA 19104; University of Manitoba, Sections of Respiratory and Critical Care Medicine, RS-317, Respiratory Hospital, 810 Sherbrooke Street, Winnipeg, Manitoba, Canada R3A 1R8

**Keywords:** Pulmonary arterial hypertension, computational model, survival

## Abstract

**Background:** Pediatric pulmonary arterial hypertension (PAH) has a long asymptomatic period with progressive vascular loss. A recent computational model of simulated PAH in humans has demonstrated that up to 70% of the pulmonary vasculature is lost before clinical PAH criteria are met. We sought to evaluate this model in pediatric subjects with PAH and evaluate whether estimated pulmonary vascular compromise (PVC) can predict survival and other clinical outcomes.

**Methods and Results:** Retrospective and prospective cohort data were collected for all subjects with PAH between 1999 and 2022 treated at our center. Cardiac catheterization and clinical data were compared with PVC estimated by the computational model. Transplant-free survival was associated with lower PVC (72% vs 88%, p<0.001). Freedom from transplant/death was also associated with a decrease in PVC over time with no significant change in PVC in subjects who died or underwent transplant. By Kaplan-Meier analysis, 10-year survival was 54% (IQR 35%, 81%) when PVC was more than 80%, compared with 100% survival (IQR 100%, 100%) when PVC was less than 80% (p<0.001). By Cox proportional hazard regression, PVC was associated with mortality (HR 1.1, p=0.008). Lower PVC was associated with better percent predicted 6-minute walk distance (-0.25, 95% CI [-0.35, -0.14], p<0.001), lower log brain natriuretic peptide (0.12, 95% CI [0.07, 0.18], p<0.001), and lower estimated 1-year mortality (0.01, 95% CI [0.01, 0.02], p<0.001).

**Conclusions:** Estimated PVC predicts transplant-free survival and other clinical outcomes in pediatric PAH and provides an adjunctive tool to potentially capture pulmonary vascular loss early in disease.

**Clinical Perspective:** *What Is New?:*

- A new computational model can estimate pulmonary vascular area loss or pulmonary vascular compromise (PVC).
- PVC can detect early vascular loss and is associated with transplant-free survival and clinical outcomes in pediatric pulmonary arterial hypertension.

*What Are the Clinical Implications?:*

- PVC may detect pulmonary vascular disease early in the disease process and could be used as an adjunct tool for management of patients with pulmonary arterial hypertension.

## INTRODUCTION

Pulmonary arterial hypertension (PAH) is characterized by aberrant remodeling in the pulmonary vasculature that contributes to a significant loss of the pulmonary vascular bed (1–6). The reduction in vascular area leads to increases in pulmonary vascular resistance (PVR) and pulmonary arterial pressures (PAP). Left untreated, PVR and PAP continue to increase leading to right heart failure and subsequently, death.

Currently, diagnosis, monitoring of treatment response, and risk prediction for PAH requires the use of invasive hemodynamic data and several functional and laboratory parameters. Cardiac catheterization provides information about PAP, pulmonary capillary wedge pressure (PCWP) or left atrial pressure (LAP), right atrial pressure (RAP), pulmonary blood flow (Qp), and PVR (2,4,7,8). New York Heart Association (NYHA) functional class, brain-type natriuretic peptide level (BNP), and 6-minute walk distance (6MW) can be used to non-invasively track progression of disease over time (7,9–13). Despite these advances in disease monitoring and the development of several treatments for PAH improving mortality in the modern era, morbidity and mortality remain high, particularly in patients with idiopathic PAH and repaired congenital heart disease (14–19). This can be explained in part by a period of progressive vascular loss early in the disease course that is asymptomatic and not associated with significant rise in PAP or PVR. In later disease, PAP and PVR begin to rise and right ventricular (RV) failure develops, leading to patient symptoms. Current strategies for monitoring patients with PAH detect disease at this later stage, when irreversible vascular loss has already occurred. Thus, risk prediction early in the disease course remains a significant challenge to improving outcomes, and adjunctive tools to monitor patients with PAH are needed to detect early disease prior to the onset of RV failure.

Computational models of the pulmonary vasculature have been previously described (20–23), and their application and limitations in modeling the pulmonary vasculature in PAH have been extensively reviewed elsewhere (24,25). To date, none of these models have been used to predict the affected area of compromised vasculature in pediatric patients with PAH. More importantly, computational modeling has yet to be a used as a variable in risk assessment. To address this, we adapted a computational model that we previously used to simulate theoretical PAH and predict pulmonary vascular loss or pulmonary vascular compromise (PVC) (26). In simulated PAH, the model demonstrated a hyperbolic relationship between PVC and mean PAP (mPAP) or PVR. In addition, it revealed that mPAP and PVR change very little until the vasculature is significantly compromised. To assess the utility of PVC in pediatric PAH, we examined a cohort of pediatric subjects with PAH at a single institution to determine whether PVC or PVC changes over time could predict transplant-free survival and other clinical outcomes in PAH independent of hemodynamic indicators of right heart failure. We hypothesized that subjects with PAH would demonstrate elevation in PVC early in disease course and that PVC would be associated with mortality and other measures of outcome in PAH.

## METHODS

### Study Population

A retrospective cohort study was performed including all subjects with a World Health Organization (WHO) Group 1 PAH diagnosis prior to 18 years of age who were between 4 to 22 years of age during the study period of 1/1/1999 to 12/31/2022. Subjects were included if they had at least two hemodynamic evaluations with right heart catheterization at our institution and had at least two years of follow-up after their first catheterization or underwent lung transplant or died prior to this timepoint. Subjects younger than 4 years of age were excluded to ensure that age-dependent pulmonary maturation was complete. Subjects with chronic lung disease were also excluded. To obtain a control population of subjects with nearly normal pulmonary vasculature, we included a random sample of patients with a WHO group 2 PH diagnosis during the study period who ultimately underwent orthotopic heart transplantation (OHT). These patients undergo serial post-transplant catheterizations and demonstrate quickly improving clinical, hemodynamic, or laboratory evidence of PH by last catheterization (27–30). De-identified data, methods, and code will be shared upon request. The study was approved by the Institutional Review Board of The Children’s Hospital of Philadelphia.

### Data Collection and Study Measures

Data was collected from an existing PAH database and from review of patient charts.

The primary exposure was PVC, and the primary outcome was transplant-free survival from the time of first catheterization.

Secondary outcomes included serum BNP, estimated 1-year mortality (as calculated by the logistic regression equation from Clabby et al. that includes RAP×PVR (31)), and 6MW (from which a percent predicted 6MW (PP6MW) (32) was calculated). Covariates included hemodynamic and other associated variables obtained at cardiac catheterization including age at catheterization, body mass index, pulmonary blood flow (Qpi), PAP, PCWP or LAP, indexed PVR (PVRi), RAP, and RAP×PVRi. Body mass index (BMI), BNP, and 6MW values within two months of each catheterization date were used. Decisions regarding clinical testing (e.g, the frequency and timing 6MW and BNP) were at the discretion of the care team. Missing data are therefore, inevitable, not likely to be missing at random. Therefore, for analyses on which these depended, case restriction was used. Imputation (or alternative strategies to address missing data) were not attempted.

### PVC Computational Model

A computational model of the normal pulmonary circulation based on studies in animals was adapted for use in humans to simulate theoretical PAH in a recently published study by Bshouty (26). This model uses PCWP, cardiac index, and mPAP to generate an estimated pulmonary vascular capacitance as a percentage of what is ideal based on estimated lung size.

From the pulmonary vascular capacitance, a percent pulmonary vascular compromise can be obtained. The methodology, algorithms, and theory behind the original model based on experimental data in dogs are detailed extensively in previous studies (33,34), and the code of the computational program can be found at https://github.com/AdamMajer/bshouty-lung-model.

Briefly, the model is an abbreviated 5-generation branching system that represents 15 generations of arterial and venous vessels bifurcating from the pulmonary artery (PA) to the left atrium (LA). Each model vessel generation represents 3 generations of real lung vessels given that distensibility is independent of diameter and equal across the pulmonary arterial tree(35–38). In addition, parent to daughter vessel length and diameters for the area and resistance calculations are based on human data from a previous study(39). The model assigns a resistance and vascular area for each branch generation under standardized conditions including a transpulmonary pressure (P_tp_) of zero and vessel transmural pressure (P_tm_) of 35 cm H_2_O. Three sets of data are entered into the computational program. General data includes tolerance or activity level for each calculation and reference point for pressure measurements that is assigned at the LA level. Patient specific data includes height, weight, lung height (P-A diameter at left atrial level), lung compliance (assumed to be normal lung compliance (C_L_) of 0.317 L/cm H_2_O), and lung volume. Hemodynamic data entered include PCWP, Qp and mPAP at P_al_ of zero and a P_pl_ of -5 cm H_2_O reflecting end-expiration. After computer iteration of a stable solution, output data includes the distribution of individual pulmonary vascular flows, arterial, capillary, and venous resistances across the whole pulmonary circulation. In addition, summary data of vascular area (PVC), mPAP, PVR, upstream (arterial), middle (capillary), and downstream (venous) resistances are generated.

### Statistical Analysis

The overall goal of the analysis was to evaluate PVC as a marker in pediatric PAH, specifically to determine whether PVC and change in PVC were associated with increased likelihood of death or transplant. Baseline characteristics of the study population were summarized via descriptive statistics. Clinical characteristics were compared using Wilcoxon rank sum and/or Fisher’s exact test between transplant-free survivors and non-transplant-free survivors at time of first and last cardiac catheterization and for all cardiac catheterizations.

Next, the change in PVC and RAPxPVC from first to last catheterization was compared among transplant-free survivors and subjects who died or underwent transplant via paired-Wilcoxon Rank Sum tests.

PVC was calculated and plotted against mPAP and PVR in a simulated model of PAH and compared with data from our population with PAH. Data from subjects with Group 1 PAH was compared with data from subjects with Group 2 PH. Lorentzian curve fitting to produce trendlines in graphs was computed using Graphpad Prism version 10.2.1 for Windows, GraphPad Software, Boston, Massachusetts, USA, www.graphpad.com.

Non-parametric Kaplan-Meier survival estimates with Log rank test were performed to evaluate differences (p-value <0.05) in median survival times between subgroups. Cut points were determined by the median of each variable from all included catheterizations. Survival hazard ratios were generated via time-varying Cox proportional hazards regression. Time-to-event was defined as the time from first catheterization to death or transplant, or to last follow-up for controls. Due to significant multicollinearity, multivariable analysis was not performed. Univariable and multivariable mixed effects linear regression models were used to evaluate the contribution of PVC and hemodynamic parameters on clinical outcomes including BNP, PP6MW, 1-year estimated mortality, and transplant-free survival while controlling for age at catheterization. One-year mortality estimates were calculated using a forward logistic regression equation from Clabby et al. (31). Log-transformed BNP was used in the analysis to ensure normality. All statistical analyses were performed using R version 4.2.0 (2022-04-22), R Foundation for Statistical Computing, Vienna, Austria, www.R-project.org.

## RESULTS

### Study Population

Fifty-eight patients met study criteria (**Table 1**). Patients were followed to a median age of 18.9 years (IQR 15.7, 22.8) with a median follow-up time from first catheterization of 10.8 years (IQR 7.5, 15.4). The median number of cardiac catheterizations per patient was 4 (IQR 3, 6). Of the 58 patients, 45 survived without transplant. Subgroups of Group 1 PAH included idiopathic PAH and congenital heart disease-related PAH. A total of 350 cardiac catheterizations were included.

**Table 1.** Baseline characteristics of patients.

| Characteristic | Lung Transplant or Death, N = 13<sup>1</sup> | Transplant-free survival, N = 45<sup>1</sup> | p-value<sup>2</sup> |
| --- | --- | --- | --- |
| Sex |  |  | >0.9 |
| Female | 9 (69%) | 29 (64%) |  |
| Male | 4 (31%) | 16 (36%) |  |
| Age at PAH Diagnosis (Years) | 6.7 (2.6, 11.1) | 4.7 (2.0, 8.0) | 0.3 |
| Age at Lung Death (Years) | 17.0 (14.1, 22.5) | NA (NA, NA) |  |

<sup>1</sup>n (%); Median (IQR)
<sup>2</sup>Fisher's exact test; Wilcoxon rank sum test

There was no difference in age at diagnosis between survivors and non-survivors.

Median age at death or transplant was 17 years. Baseline patient characteristics at first cardiac catheterization at our institution are presented in **Table 2**. Non-survivors were older at first catheterization. They also had lower PP6MW and higher BNP, mPAP, PCWP/left atrial pressure (LAP), indexed PVR (PVRi), right atrial pressure (RAP), and RAPxPVRi. Similar trends were seen when evaluating data from patients’ most recent catheterization or last catheterization prior to lung transplant or death. In addition, non-survivors were older at the time of the last cardiac catheterization (16.7 vs. 13.6 years, p=0.02) (**Table S1**). When evaluating data from all cardiac catheterizations combined, similar hemodynamic trends were again demonstrated; non-survivors had significantly higher body mass index (18.0 vs. 16.9 kg/m2, p=0.047) and lower Qpi (3.28 vs 3.87 L/min/m2, p=0.001) (**Table S2**).

**Table 2.** Comparison of data at first catheterization from PAH patients with and without transplant-free survival.

| Characteristic | N | Lung Transplant or Death, N = 13<sup>1</sup> | Transplant-free survival, N = 45<sup>1</sup> | p-value<sup>2</sup> |
| --- | --- | --- | --- | --- |
| Age at Cath | 58 | 12.7 (7.6, 14.3) | 6.4 (4.9, 8.5) | 0.012 |
| BMI | 58 | 16.85 (15.31, 17.57) | 15.54 (14.46, 17.84) | 0.6 |
| BNP | 38 | 324 (203, 1,326) | 60 (30, 135) | 0.010 |
| % Pred. 6MW | 32 | 42 (33, 59) | 69 (56, 80) | 0.031 |
| QPi | 57 | 3.65 (2.55, 5.37) | 3.90 (3.30, 4.87) | 0.6 |
| mPAP | 57 | 64 (62, 78) | 34 (25, 55) | <0.001 |
| mPCWP | 56 | 12.5 (12.0, 15.0) | 10.0 (9.0, 12.2) | 0.007 |
| PVRi | 57 | 17 (9, 24) | 6 (3, 10) | 0.002 |
| RAP | 57 | 10.0 (7.8, 11.0) | 7.0 (5.0, 9.0) | <0.001 |
| RAPxPVRi | 57 | 184 (66, 255) | 32 (19, 68) | <0.001 |

<sup>1</sup>Median (IQR)
<sup>2</sup>Wilcoxon rank sum test

### PVC modeling in pediatric PAH

In our previous report, at a fixed LAP of 7.5 mmHg, stepwise increases of mPAP and PVR demonstrated a curvilinear relationship with PVC (26) (**Figures 1A** and **2A**, respectively). In addition, the PVC vs. PVR curve shifts downward and to the right with increasing Qp, and the PVC vs. mPAP curve shifts upward and to the left with increasing Qp.

**Figure 1.**
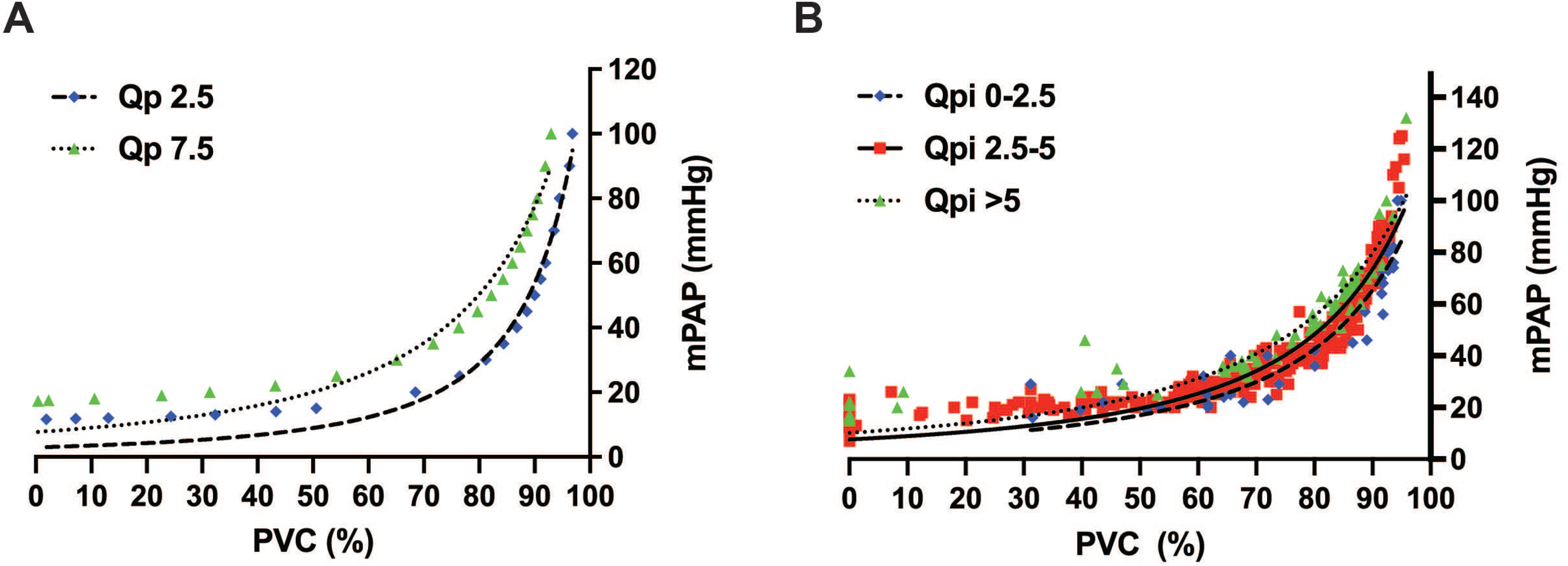
Computed pulmonary vascular compromise (PVC) versus changes in mean pulmonary artery pressure (mPAP) in simulated pulmonary arterial hypertension (PAH) and in a pediatric PAH patient population. (*A*) PVC in simulated PAH calculated using a stepwise increase in mPAP in mmHg and either a pulmonary blood flow (Qp) of 2.5 L/min or 7.5 L/min. (*B*) PVC in a pediatric PAH study group calculated similarly and divided into three groups of indexed pulmonary blood flow (Qpi) in L/min/m2. Lorentzian curve fitting used to generate trendlines.

**Figure 2.**
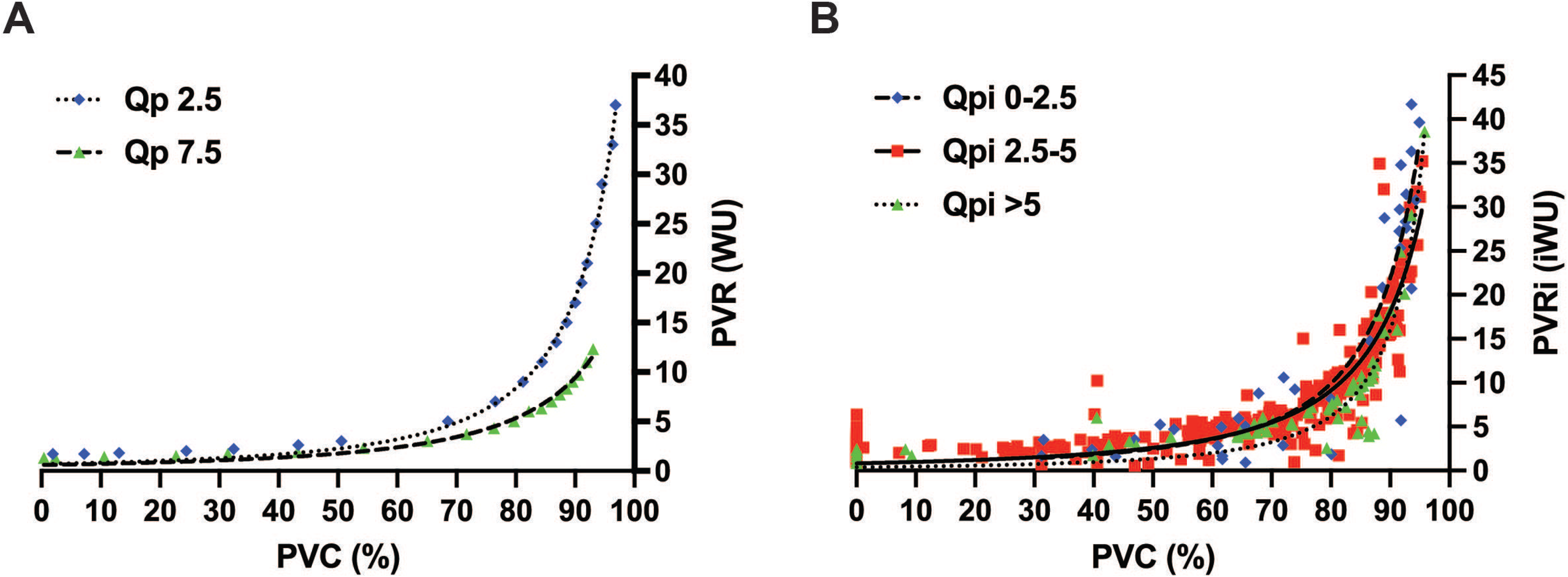
Computed PVC versus changed in indexed pulmonary vascular resistance (PVRi) in simulated PAH and in a pediatric PAH patient population. (*A*) PVC in simulated PAH calculated using a stepwise increase in PVR in Wood units and either a pulmonary blood flow (Qp) of 2.5 L/min or 7.5 L/min. (*B*) PVC versus PVRi in pediatric PAH study group calculated similarly and divided into three groups of indexed pulmonary blood flow (Qpi) in L/min/m2. Lorentzian curve fitting used to generate trendlines.

To evaluate PVC in actual pediatric patients, data from two patient populations were assessed: 350 cardiac catheterizations from 58 patients with documented Group 1 PAH and 75 cardiac catheterizations from eight control Group 2 PH patients who had undergone OHT with eventual resolution of PH on catheterization. Baseline characteristics and cardiac catheterization data for control patients are shown in **Tables S3-S6**. First, PVC for all patients was plotted vs. either mPAP (**Figure 1B**) or PVRi (**Figure 2B**); the relationship was similar to the simulated PAH model. Using Lorentzian curve fitting to predict Qpi trendlines, we saw a curvilinear relationship that shifted the curves with increased cardiac flow. There were no significant changes in patient mPAP or PVRi until the PVC was more than 70%. Thereafter, mPAP and PVRi rose rapidly with increasing PVC.

There were discrete differences in PVC when comparing patients with PAH and patients post-OHT. In PAH, PVC increased with rising mPAP and PVRi (**Figures 3A** and **3B**, blue diamonds). Data from post-OHT patients revealed normal mPAP and PVRi and PVC ranging from zero to 80% (**Figures 3A** and **3B**, red squares). No post-OHT patient had PVC greater than 80%. All post-OHT patients with any PVC early after transplantation improved to zero or nearly zero in a periodic but decreasing fashion by the end of our data collection period, as illustrated in an example patient included in **Figure 4**.

**Figure 3.**
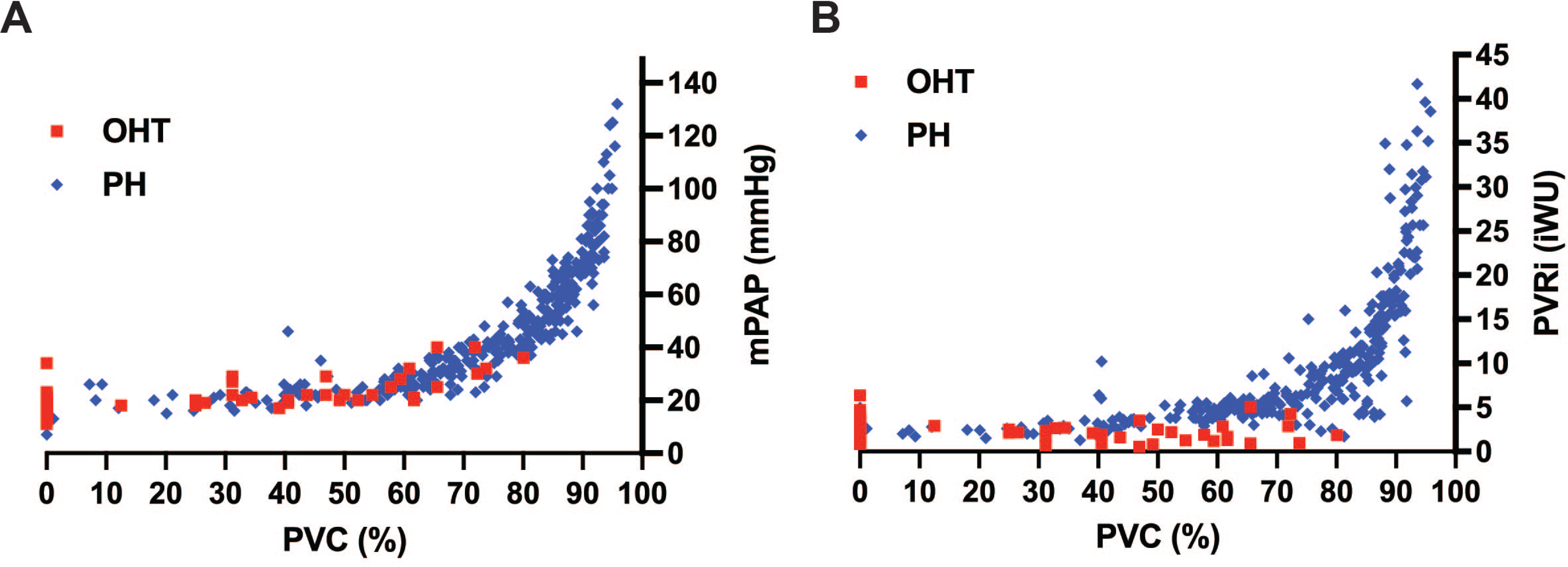
Computed PVC in pediatric patients with PAH and patients with WHO Group 2 pulmonary hypertension. (*A*) PVC vs. mPAP. (*B*) PVC vs. PVRi.

**Figure 4.**
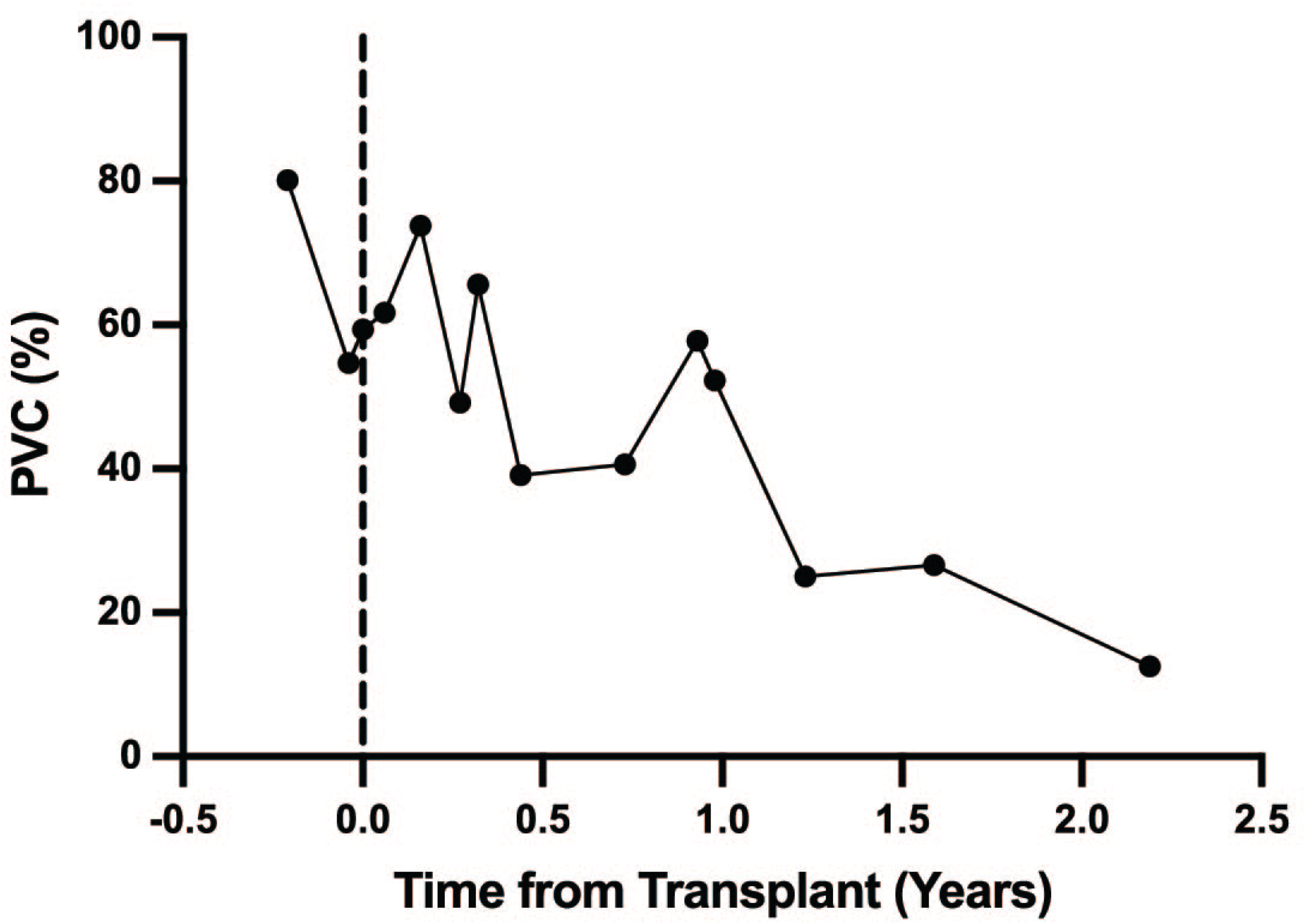
Improvement in pulmonary vascular compromise over time following orthotopic heart transplantation in a pediatric patient with WHO group 2 pulmonary hypertension.

In transplant-free survivors, PVC at last catheterization was significantly lower than at first catheterization (66% vs. 77%, p=0.02). In non-survivors no difference in PVC was seen between first and last catheterization (88% vs. 88%, p=0.94). PVC in transplant-free survivors was lower than PVC in non-survivors at both first and last catheterization (p<0.001 for both, **Table 3**). Similar trends were demonstrated when comparing right ventricular workload responses as estimated by RAP×PVR in patients with (26 vs. 44, p=0.001) and without transplant-free survival (108 vs. 119, p=0.79).

**Table 3.** Comparison of PVC and RAP × PVC at time of first, last, and all catheterizations in PAH patients with and without transplant-free survival.

| Catheterization | Characteristic | N | Lung transplant or Death, N = 13<sup>1</sup> | Transplant-free survival, N = 45<sup>1</sup> | p-value<sup>2</sup> |
| --- | --- | --- | --- | --- | --- |
| First |  |  |  |  |  |
|  | PVC | 52 | 88 (85, 92) | 77 (56, 84) | <0.001 |
|  | RAPxPVC | 52 | 818 (617, 982) | 445 (258, 583) | <0.001 |
| Last |  |  |  |  |  |
|  | PVC | 58 | 88 (87, 92) | 66 (46, 77) | <0.001 |
|  | RAPxPVC | 56 | 731 (576, 1,544) | 363 (251, 557) | <0.001 |
| All |  |  |  |  |  |
|  | PVC | 265 | 88 (85, 92) | 72 (58, 84) | <0.001 |
|  | RAPxPVC | 259 | 748 (580, 959) | 441 (294, 584) | <0.001 |

<sup>1</sup>Median (IQR)
<sup>2</sup>Wilcoxon rank sum test

### PVC and Associated Factors as Predictors of Survival

Kaplan-Meier survival curves were generated for PVC, PVRi, mPAP, RAP×PVC, BNP, and PP6MW using data from the first (**Figure S1**) and most recent cardiac catheterization (**Figure 5**). Starting from the most recent catheterization, 10-year survival was estimated at 100% when PVC was less than 80%, compared to 54% survival when PVC was higher than 80% (p< 0.001) (**Figure 5A**). Similarly, a PVRi threshold of 8 iWU and a mPAP threshold of 42 mmHg correlated with difference in 10-year survival [100% vs. 61% for PVRi (p<0.001) and 100% vs. 60% for mPAP (p<0.001)] (**Figures 5B** and **5C**). Ten-year survival estimates also differed with an RAPxPVC threshold of 510 [97% vs. 66% (p=0.002)], BNP threshold of 65 pg/ml [94% vs. 68% (p=0.036)] and PP6MW threshold of 72% [100% vs. 59%] (p<0.001)] (**Figures 5D-F**).

**Figure 5.**
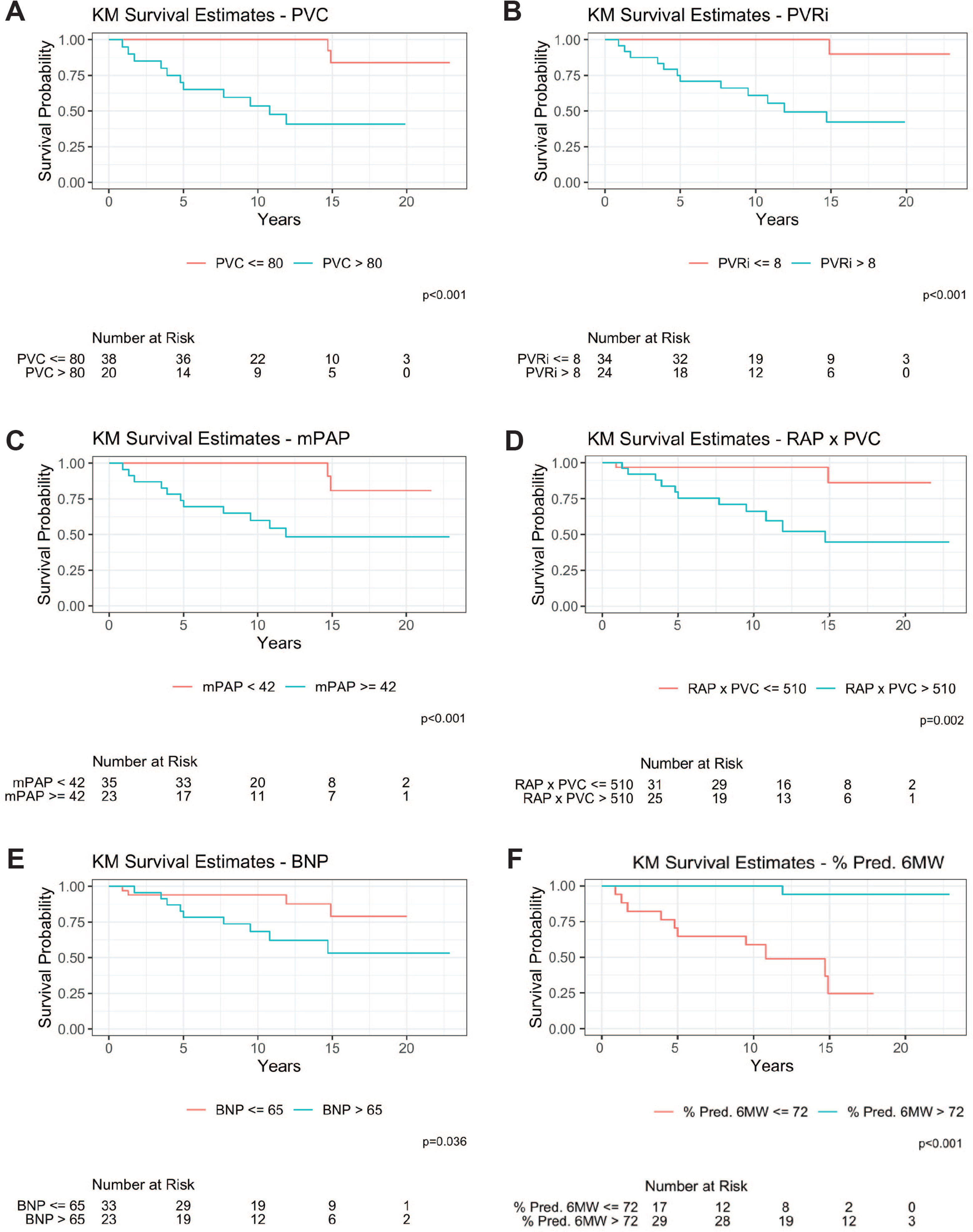
Kaplan-Meier survival estimates for transplant-free survival at last cath. (*A*) 10-year survival for PVC > 80% is 54% (95% CI 35%, 81%), *p* < 0.001. (*B*) 10-year survival for PVRi > 8 iWU is 61% (95% CI 44%, 85%), *p*< 0.001. (*C*) 10-year survival for mPAP ≥ 42 mmHg is 60% (95% CI 43%, 84%), *p*<0.001. (*D*) 10-year survival for RAPxPVC > 510 is 66% (95% CI 49%, 88%), *p*=0.002. (*E*) 10-year survival for BNP > 65 pg/mL is 68% (95% CI 51%, 91%), *p*=0.036. (*F*) 10-year survival for percent predicted 6-minute walk distance ≤ 72% is 59% (95% CI 40%, 88%), *p*<0.001. Lung transplant or death (n = 13) and transplant-free survival (n = 45).

Mortality was not associated with age at diagnosis or Qpi (**Table 4**). Age at catheterization, BNP, PP6MW, RAP, LAP, mPAP, PVRi, RAPxPVRi, PVC, and RAPxPVC all predicted mortality with statistical significance. Hazard ratios were close to 1 for most variables. The most impactful predictors of mortality were age at cardiac catheterization (HR 1.19), RAP (HR 1.22), and Qpi (HR 0.61, but p=0.092).

**Table 4.** Univariable cox regression with time varying covariates to evaluate effects of variables on time to survival.

| Characteristic | N | HR<sup>1</sup> | 95% CI<sup>1</sup> | p-value |
| --- | --- | --- | --- | --- |
| Diagnosis Age | 274 | 1.07 | 0.95, 1.20 | 0.3 |
| Age at Catheterization | 274 | 1.19 | 1.02, 1.38 | 0.024 |
| BNP | 237 | 1.00 | 1.00, 1.00 | <0.001 |
| % Pred. 6MW | 202 | 0.95 | 0.91, 0.99 | 0.010 |
| RAP | 267 | 1.22 | 1.13, 1.32 | <0.001 |
| PAP | 272 | 1.03 | 1.01, 1.05 | 0.001 |
| LAP | 271 | 1.09 | 1.01, 1.19 | 0.029 |
| QPi | 273 | 0.61 | 0.34, 1.08 | 0.092 |
| PVRi | 273 | 1.11 | 1.05, 1.18 | <0.001 |
| RAP x PVRi | 267 | 1.00 | 1.00, 1.01 | <0.001 |
| PVC | 265 | 1.10 | 1.02, 1.18 | 0.008 |
| RAP x PVC | 259 | 1.00 | 1.00, 1.00 | <0.001 |

<sup>1</sup>HR = Hazard Ratio, CI = Confidence Interval

### PVC as a Prognostic Factor in Additional Clinical Outcomes

Univariate mixed model analysis demonstrated that all variables except Qpi are significant predictors of PP6MW, BNP, and estimated 1-year mortality by the Clabby et al regression equation in our pediatric PAH population (**Table 5**). In multivariate mixed models, PVRi, RAP, PP6MW, and log Qpi were associated with estimated 1-year mortality. When PVRi was replaced with PVC, only PVC, RAP, and log Qpi were significant predictors of estimated 1-year mortality (**Table 6**).

**Table 5.**
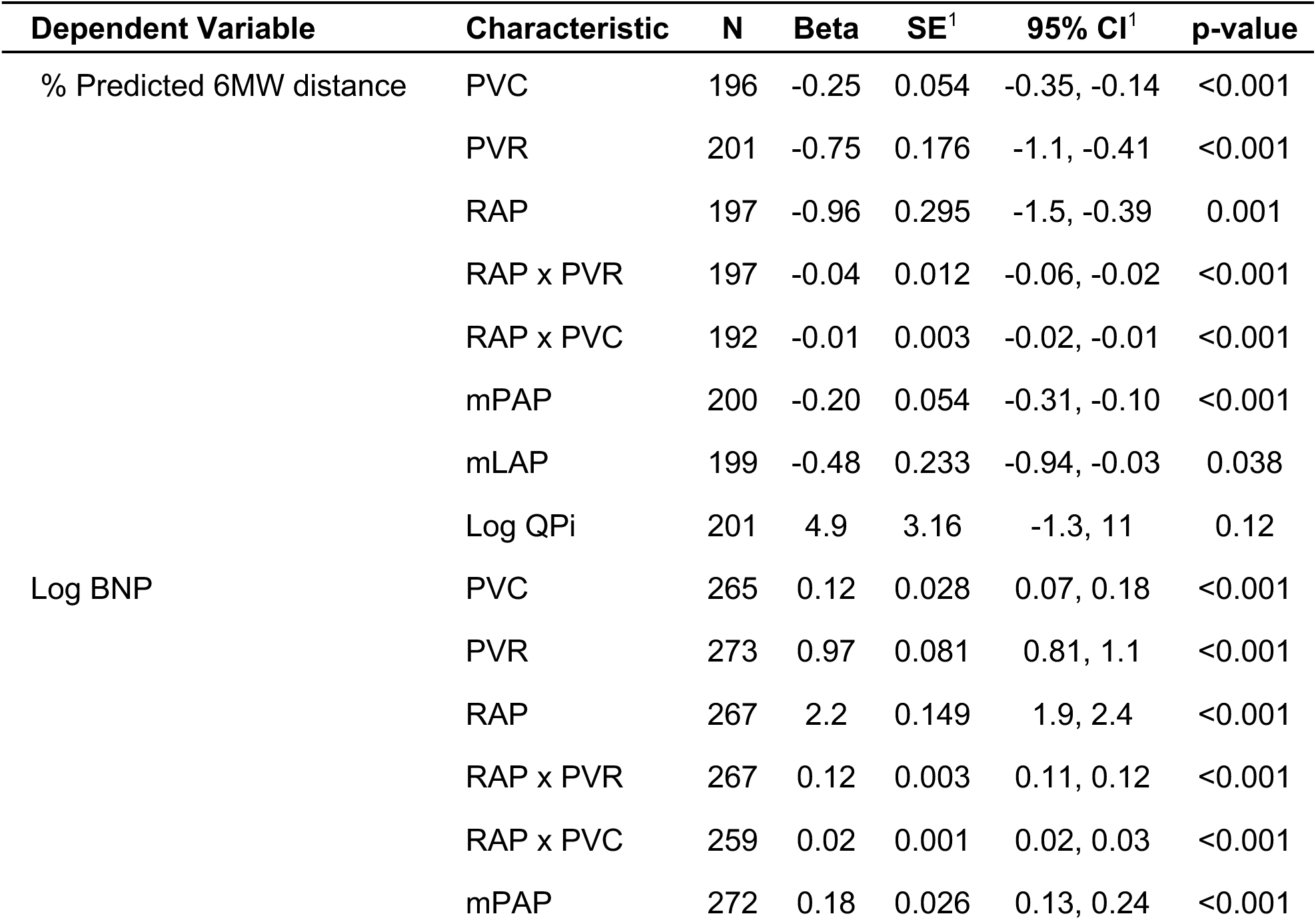
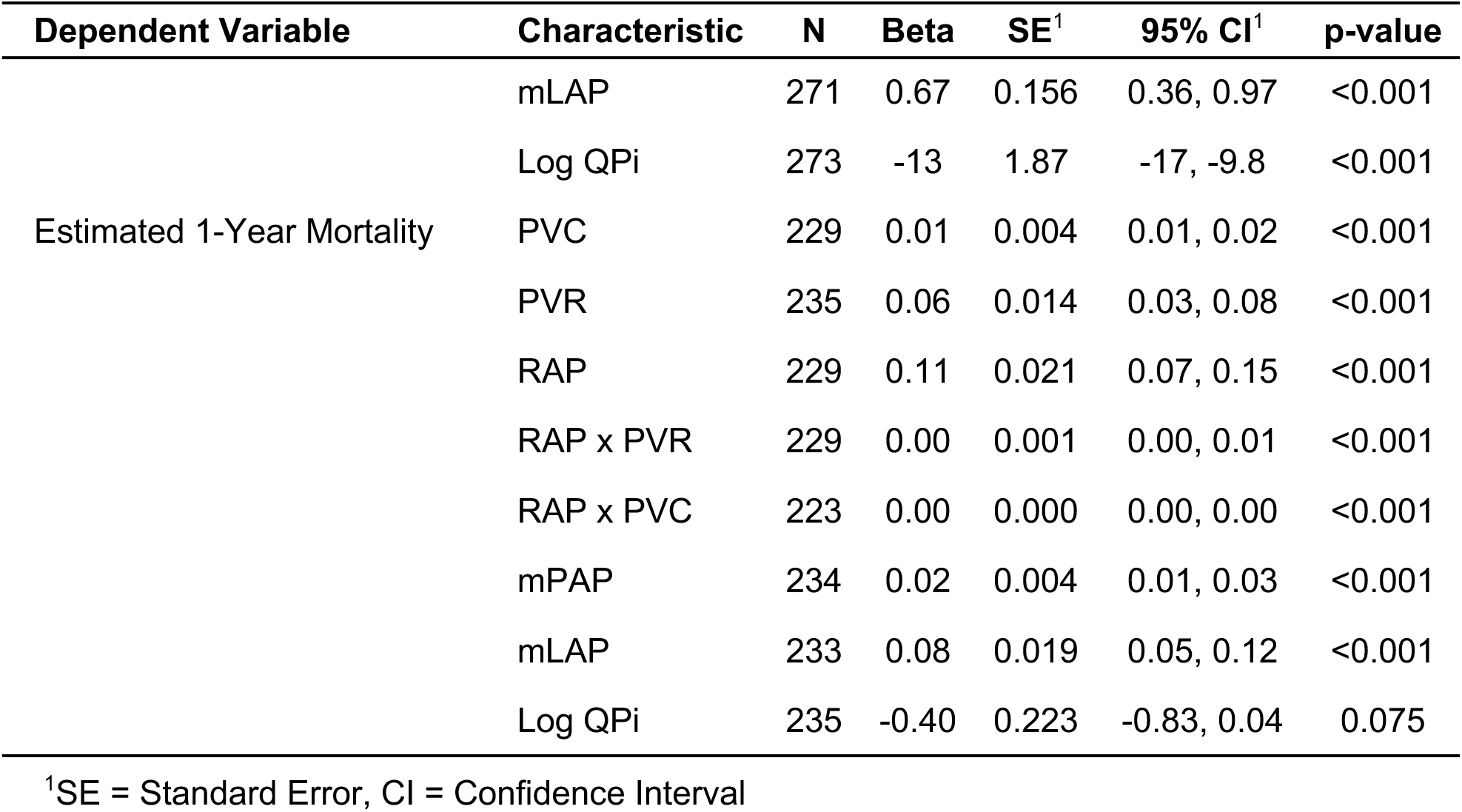
Univariate mixed model analysis of predictors of 6MW, BNP, and estimated 1-year mortality.

**Table 6.** Multivariable mixed model analyses of predictors of 6MW, BNP, and estimated 1-year mortality Variable.

| Dependent Variable | Model | Characteristic | Beta | SE<sup>1</sup> | 95% CI<sup>1</sup> | p-value |
| --- | --- | --- | --- | --- | --- | --- |
| Estimated 1-year mortality | 12 | PVR | 0.78 | 0.093 | 0.60, 0.96 | <0.001 |
|  | 12 | RAP | 1.9 | 0.189 | 1.6, 2.3 | <0.001 |
|  | 12 | % Pred. 6MW | 0.12 | 0.039 | 0.04, 0.19 | 0.003 |
|  | 12 | Log BNP | -0.40 | 0.445 | -1.3, 0.47 | 0.4 |
|  | 12 | Log QPi | -4.1 | 1.47 | -7.0, -1.2 | 0.005 |
| Estimated 1-year mortality | 13 | PVC | 0.08 | 0.033 | 0.01, 0.14 | 0.017 |
|  | 13 | RAP | 2.3 | 0.218 | 1.8, 2.7 | <0.001 |
|  | 13 | % Pred. 6MW | 0.06 | 0.047 | -0.04, 0.15 | 0.2 |
|  | 13 | Log BNP | -0.03 | 0.548 | -1.1, 1.0 | >0.9 |
|  | 13 | Log QPi | -6.9 | 1.80 | -10, -3.3 | <0.001 |

<sup>1</sup>SE = Standard Error, CI = Confidence Interval

## DISCUSSION

In this single center retrospective cohort study, we demonstrated that a computational model can be applied to quantify progressive pulmonary vascular loss in pediatric patients with PAH, and that worsening PVC was consistently associated with worse transplant-free survival. Consistent with an estimate in a theoretical PAH model by Bshouty (26), PVC greater than 70% indicated severe PAH. Furthermore, PVC was significantly associated with mortality in pediatric PAH. Given this data, the model’s simple interface, and iterations taking seconds to predict PVC, any clinician with patient hemodynamic data can employ this adjunctive diagnostic tool for PAH.

This addresses a critical challenge in the chronic treatment of young patients with PAH. PAH is a silent and severe disease. Early detection is difficult as patients with mild disease are often asymptomatic or present with nonspecific symptoms. Diagnosis is often delayed for years, during which vascular remodeling causes lasting and sometimes irreversible damage to the vascular bed (5,18,40,41). The results of the current study may provide a pathophysiologic explanation for this early asymptomatic period. In our model, mPAP and PVRi change very little until 70% of the pulmonary vascular bed is lost. After crossing the 70% threshold of PVC, both mPAP and PVRi rapidly rise, leading to RV hypertension and ultimately symptomatic RV failure. Current World Symposium of PH diagnostic criteria for PAH requires mPAP greater than 20 mmHg, PVRi greater than 3 iWU, and PCPW or LAP less than 15 mmHg. While these criteria identify patients with significant PAH, there is likely a distinct group of patients with early disease, mild disease, or concurrent left-sided disease who are excluded from the definition.

The model presented here provides an adjunctive diagnostic tool to identify PAH at an earlier stage and in patients with concurrent left-sided heart disease.

PAH is marked by both fixed and reversible vaso-occlusive lesions that worsen over time. As the disease progresses, the area of functional pulmonary vasculature is reduced. These phenomena have been reported in earlier studies examining biopsies of affected lungs (2,4,12,42). Recent advances in computed tomography (CT) and magnetic resonance imaging (MRI) have provided some insight into this process as well (43,44). MRI-based pulmonary blood flow analysis can measure perfusion changes in PAH and infers that there is a reduction in the functional pulmonary vasculature (43,45). However, these imaging modalities are limited in the resolution to delineate distal pre-capillary arterioles. Computational modeling to estimate vascular area can provide a reasonable alternative until improved resolution in CT or MRI develops. While lung biopsy can provide information about the degree of disease in distal arterioles, it is significantly more invasive than computational modeling, and detection of patchy disease is difficult due to sampling limitations.

The current study demonstrated that PVC and RAP×PVC are significant predictors of clinical variables such as BNP and PP6MW. Both BNP and PP6MW have been used to monitor clinical response to PAH therapy in multiple small clinical trials (46–51). There is also some evidence that BNP correlates with WHO Functional Class in children with PH (52), and a BNP less than 50 pg/ml is associated with improved survival in children with PAH (14). While BNP and RAP are reflective of a failing RV and increased mortality risk, PVC and the hemodynamic variables required to calculate it are independent of RV function. Thus, PVC is a potential adjunctive tool for the evaluation of PAH.

In this study, PVC and RAP×PVC were associated with both estimated and actual mortality. PVC and RAP×PVC were associated with estimated 1-year mortality as calculated by the logistic regression equation from Clabby et al (31). Although most studies on predictors of mortality are derived from databases composed predominantly of adult patients with PH, there have been several studies examining risk factors for children (14,16–19,53,54). Early studies in pediatric PH identified elevated mean RAP and a decreased stroke volume index as risk factors for increased mortality. Several more recent studies of PH registries reported poor growth, younger age, worse WHO functional class, elevated BNP, elevated uric acid, lower Qp, lower systolic blood pressure, and a higher ratio of mPAP to mean systemic arterial pressure (9,13,17,18,53) as additional risk factors. Data from the REVEAL registry confirmed several of these variables including age at diagnosis, lower Qp, higher PVR, and poor response to acute vasoreactivity testing (14,55). PVC is an additional diagnostic tool to help monitor progression of the disease and potentially predict mortality.

Median PVC in transplant-free survivors was lower at the time of the last cardiac catheterization compared with the first catheterization. Conversely, PVC did not improve in patients with lung transplant or death. This pattern mirrors that of RAPxPVR, a metric known to be associated with survival in patients with PH. These findings may suggest that transplant-free survivors had a therapeutic response to PH treatment and were able to recruit additional functional pulmonary vasculature, while non-survivors may have had more advanced disease at diagnosis with limited vascular reserve. Thus, serial PVC measurements may be able to detect subtle improvements in vascular capacity over time and could potentially be used to monitor response to treatment.

There are multiple limitations to this study. As in the previous study by Bshouty, multiple assumptions were made that may make use of the model in other types of PH difficult. First, we assume that the lung compliance is normal for all patients, but lung compliance might be different among different pediatric patient populations. For instance, patients with bronchopulmonary dysplasia (BPD) have severely reduced lung compliance both early in life and sometimes into adulthood (56,57). We also assumed that the vascular growth of our patient population was normal. Early studies have indicated that pulmonary vascular growth and remodeling continues into childhood (58), and it is reasonable to surmise that development of pulmonary vasculature may be an ongoing process in our patients. In BPD, pulmonary vascular development can be deranged, decreasing the amount of vascular bed area (59). In this study, we excluded patients with BPD and patients under the age of four years to reduce variability in pulmonary vascular area. Future studies to examine PVC in infants and patients with BPD are warranted to determine whether this computational model correlates with hemodynamic parameters in these populations.

An additional limitation is our inclusion of patients only after data from the first cardiac catheterization is available. This excludes patients who died or underwent lung transplant prior to obtaining a cardiac catheterization, which leads to immortal time bias. Furthermore, descriptive statistics were included at time of first and last cardiac catheterization. As different patients underwent cardiac catheterizations at different times in the disease course, these comparisons do not account for the timing of the catheterization. However, survival analysis was performed later in the study using time-varying cox regression analysis, which does account for timing of cardiac catheterization.

This study was limited to assessment of PAH in children at a single institution. Our data are predominantly derived from children with Group 1 PAH including congenital heart disease-associated PAH and idiopathic PAH. Adult PAH populations are enriched with idiopathic PAH and connective tissue disorder-associated PAH (60). Whether the model will work similarly in other pediatric and adult PAH populations is unknown and warrants future studies. In addition, we will make the model program publicly available upon publication, and additional institutions will be able to assess PAH and independently assess our model.

Finally, PVC is an adjunctive tool and should not supplant other predictors and diagnostics of PAH and heart failure including PVR, mPAP, RAP, and BNP. PVC was found to be closely associated with many variables, and whether it is a better independent predictor cannot be ascertained. Nevertheless, we do believe that this adjunctive tool has the potential to improve early diagnosis and monitoring of progression of PAH.

## CONCLUSION

The computational model presented in this work used hemodynamic data and lung dimensions from pediatric patients with PAH to generate values of PVC that conformed to theoretically modeled PAH. PVC accurately predicted clinical outcomes in PAH similar to all current predictors. Future studies are needed to confirm these observations in other patient populations and to evaluate whether PVC decreases with PH-directed therapy.

## Data Availability

All data will be publicly available from the corresponding author upon publication. The code for building the computational model can be found at https://github.com/AdamMajer/bshouty-lung-model.

## Acknowledgements

We would like to thank the Ayla Gunner Prushansky Research Fund for support of this manuscript.

